# Diagnostic accuracy of RT-PCR for detection of SARS-CoV-2 compared to a “composite reference standard” in hospitalized patients

**DOI:** 10.1101/2021.02.18.21252016

**Authors:** Noah Reich, Christopher F. Lowe, David Puddicombe, Nancy Matic, Jesse Greiner, Janet Simons, Victor Leung, Terry Chu, Hiten Naik, Nick Myles, Laura Burns, Marc G. Romney, Gordon Ritchie, Sylvie Champagne, Kent Dooley, Inna Sekirov, Aleksandra Stefanovic

**Affiliations:** Faculty of Medicine, University of British Columbia, Canada; Division of Medical Microbiology and Virology, Providence Health Care, St. Paul’s Hospital; Department of Pathology and Laboratory Medicine, University of British Columbia, Vancouver, Canada; Department of Medicine, Division of General Internal Medicine, University of British Columbia, Vancouver, Canada; Perinatal Services BC, Provincial Health Service Authority, Vancouver, BC; Life Labs, Vancouver, Canada; British Columbia Center for Disease Control, Vancouver, BC

**Author notes:** Corresponding Author, Aleksandra Stefanovic MD, FRCPC, St. Paul’s Hospital, Providence Health Care, Virology Laboratory, 1081 Burrard St. Vancouver, BC, Canada, V6Z 1Y6.

**Keywords:** Coronavirus, COVID-19, SARS-CoV-2, diagnostic accuracy, sensitivity, serology

## Abstract

**Background:** COVID-19 caused by the novel coronavirus SARS-CoV-2 has caused the greatest public health emergency of our time. Accurate laboratory detection of the virus is critical in order to contain the spread. Although real-time polymerase chain reaction (PCR) has been the cornerstone of laboratory diagnosis, there have been conflicting reports on the diagnostic accuracy of this method.

**Methods:** A retrospective review was performed on all hospitalized patients tested for SARS-CoV-2 (at St. Pauls Hospital in Vancouver, BC) from March 13 – April 12, 2020. Diagnostic accuracy of initial PCR on nasopharyngeal (NP) swabs was determined against a composite reference standard which included a clinical assessment of the likelihood of COVID-19 by medical experts, initial and repeat PCR, and post-hoc serological testing.

**Results:** A total of 323 patients were included in the study, 33 (10.2%) tested positive and 290 (89.8%) tested negative by initial PCR. Patients testing positive were more likely to exhibit features of cough (66.7% vs 39.3%), shortness of breath (63.6% vs 35.9%), fever (72.7% vs 27.6%), radiographic findings (83.3% vs 39.6%) and severe outcomes including ICU admission (24.2% vs 9.7%) and mortality (21.2% vs 6.2%) compared to patients testing negative. Serology was performed on 90 patients and correlation between serology and PCR was 98.9%. There were 90 patients included in the composite reference standard. Compared to the composite reference standard, initial PCR had sensitivity of 94.7% (95% CI 74.0 to 99.9%), specificity of 100% (95% CI 94.9 to 100%), positive predictive value of 100% (95% CI 81.5 to 100%) and a negative predictive value of 98.6% (95% CI 92.5 to 100%).

**Discussion:** Our study showed high sensitivity of PCR on NP swab specimens when compared to composite reference standard in hospitalized patients. High correlation of PCR with serological testing further increased confidence in the diagnostic reliability of properly collected NP swabs.

## Introduction

COVID-19, a disease caused by the novel coronavirus virus SARS-CoV-2, has caused an unprecedented global pandemic and rapidly developed into the greatest public health emergency of our time [1] [2].The number of COVID-19 cases has reached over 100,000,000 and resulted in more than 2,000,000 deaths globally at the time of this manuscript [3]. Accurate laboratory detection has been paramount to controlling SARS-CoV-2 spread through identification of infected individuals and areas of outbreak in the community and healthcare facilities. Furthermore, expeditious testing of infected individuals has been critical to the public health response of rapid case identification, contact tracing and quarantine [4].

Clinical manifestations of COVID-19 range from asymptomatic or mild infections to severe pneumonia, respiratory failure and death [5]. While most cases of COVID-19 are mild, up to 20% require hospitalization and 5% intensive care [6] [7]. Hospitalized patients most commonly present with fever, cough, shortness of breath, myalgia, fatigue, lymphopenia and bilateral ground glass opacities on chest imaging. [6] [7] [8]

Molecular methods such as PCR are the mainstay for the diagnosis of COVID-19 in hospitalized patients. Significant differences in molecular assays exist with respect to extraction methods, target detection, specimen validation, instrumentation, and analytical sensitivity. The SARS-CoV-2 genome has been well characterized allowing for primer development targeting different components of the viral genome [9]. Commonly used primer targets include RdRP gene (RNA-dependent RNA polymerase), E gene (envelope glycoprotein), N gene (nucleocapsid phosphoprotein), ORF 1a/b genes (open reading frame), and S gene (spike glycoprotein). As part of the *in vitro* diagnostic validation and regulatory approval, molecular assays undergo assessment of analytic sensitivity and specificity; however, clinical diagnostic performance is less well described. Reports of initial false negative SARS-CoV-2 results by PCR, later diagnosed with COVID-19 by chest CT and repeat PCR, have emerged [10] [11]. As SARS-CoV-2 serological testing has become available, this has presented another testing modality to retrospectively adjudicate suspected cases with negative PCR [12]. The sensitivity and specificity of serology are estimated in the range of 90-100% and 95-100%, respectively [13].

As no single reference standard for diagnosing COVID-19 exists, we aim to determine the clinical sensitivity, specificity, positive and negative predictive values of a SARS-CoV-2 PCR assay in acutely ill patients admitted to hospital using a composite reference method including clinical assessment, molecular testing and serology for SARS-CoV-2.

## Methods

### Participant selection

All adult patients aged ≥18 years admitted to an acute care hospital for >24 hrs tested by PCR for SARS-CoV-2 from March 13 to April 17, 2020 were included in the study. Patients were excluded from the study if they had testing performed >40 days after symptom onset [14].

### Data collection

Data were collected retrospectively from the hospital’s electronic medical record system. Patient information included age, gender, medical comorbidities, symptoms, vital signs, laboratory and imaging findings at the time of presentation to hospital. Presence of keywords on imaging reports typical of COVID-19 infection (i.e., ground-glass, patchy infiltrates, opacification, airspace disease, consolidation, crazy paving sign) as reported in the literature, were used as a dichotomous variable [11] [15]. Admission date, discharge date, ICU admission, and mortality were also collected. This study was approved by the Research Ethics Board, University of British Columbia.

### Diagnostic testing

Specimens for testing included nasopharyngeal swab, sputum, tracheal aspirate, or bronchoalveolar lavage. Testing for viral RNA consisted of one of two commercial methods: LightMix® Real-Time PCR COVID-19 assay for the Envelope E-gene (TIB Molbiol, Germany) with amplification on the Roche LightCycler® 480, or Roche cobas® SARS-CoV-2 Qualitative Assay on the cobas® 6800 System for detection of ORF1/a and E-genes (Roche Molecular Diagnostics, Laval, QC). Antibody testing was performed on patients with serum collected ≥2 weeks and <4 months after PCR testing or symptom onset. Elecsys® Anti-SARS-CoV-2 assay (Roche) using recombinant protein representing the nucleocapsid (N) antigen for determination of total antibodies was performed on the Roche cobas® e601. The test result is given as a cut-off index (COI) with COI≥1.0 considered “reactive” and COI<1.0 as “non-reactive” as per package insert [16].

### Assessment of clinical likelihood of COVID-19

Patients were classified as having low, moderate, or high likelihood of COVID-19 based on chart review and clinical assessment [6] [7] [8]. The assessors were asked to review epidemiologic risk factors, clinical signs and symptoms, imaging findings, and laboratory results (other than SARS-CoV-2 PCR or serology results) to judge the probability of COVID-19 infection. Low probability cases had a clear alternative diagnosis explaining the clinical presentation and/or clinical features inconsistent with a COVID-19 infection; moderate probability cases had compatible clinical features and/or radiology findings but a presumptive alternative diagnosis; and high probability cases presented with compatible clinical features, radiological findings, no alternative diagnosis and/or an epidemiologic link to a known COVID-19 case. Patients deemed moderate to high risk for COVID-19 on initial assessment, underwent further review by an “expert panel” consisting of two internal medicine specialists caring for patients on dedicated COVID-19 hospital units. Any discordance in “expert” assessment was reviewed by an infectious disease specialist. Reviewers were not blinded to the PCR test result as it was reported in the electronic medical chart but were blinded to the serological result.

### Composite reference standard

The composite reference standard included the clinical assessment, any PCR result and serology. Positive reference standard was defined as testing positive on at least 2 out of 3 modalities (moderate or high clinical likelihood, PCR and serology). Composite reference testing was only performed on cases which had all 3 modalities, including serology, available. Cases with no serology available but PCR conversion from negative to positive for the same clinical episode within 4 weeks were deemed false negative on initial PCR and were included in the calculation of diagnostic accuracy. PCR reversion from positive to negative in patients with resolved clinical symptoms within the follow-up period was not considered false positive as such conversion was considered as part of the natural history of the disease.

### Statistical Analysis

The analysis population included all eligible patients who were admitted to hospital during the study period. Pearson’s chi-squared test was used to measure the association between binary or categorical variables and PCR result. T-tests were performed to compare the equality of means between continuous variables. For the sensitivity, specificity, positive and negative predictive values, exact binomial 95% confidence intervals were calculated. Stata version 14.1 was used for all analyses [17].

## Results

### Epidemiological data

There were 323 patients included in the study of which 33 (10.2%) tested positive and 290 (89.8%) tested negative on initial PCR. The mean age of the PCR positive and negative cases was 70.4 and 58.3, respectively (Table 1). Males and females comprised 69.3% and 30.3% of PCR negative cases and 66.7% and 33.3% of PCR positive cases, respectively. The average time from symptom onset to hospital admission in PCR positive and negative cases was 4.8 and 4.2 days, respectively. Of the PCR positive cases, 30.3% reported having a high risk exposure to a COVID-19 confirmed case and 12.1% reported a travel history, while only 1.7% of PCR negative cases had a high risk exposure and 3.1% had a travel history.

**Table 1.** Clinical Characteristics of the Study Patients According to PCR result.

| Characteristic | PCR positive<br>(N=33) | PCR negative<br>(N=290) | p-value |
| --- | --- | --- | --- |
| **Age**<br>Mean (SD) | 70.4 (17.8) | 58.3 (17.5) | <0.001 |
| **Gender**<br>Male – no. (%)<br>Female – no. (%)<br>Undifferentiated – no. (%) | 22 (66.7)<br>11 (33.3)<br>0 | 201 (69.3%)<br>88 (30.3)<br>1 (0.3) | 0.891 |
| **Exposure to known source of transmission within past 14 days** – no. (%) | 10 (30.3) | 5 (1.7) | <0.001 |
| **Travel history outside of Canada** | 4 (12.1) | 9 (3.1) | 0.01 |
| **Days from symptom onset to admission to hospital**<br>Mean (SD) | 4.8 (3.2) | 4.2 (6.1) | 0.31 |
| **Symptoms on admission** – no. (%)<br>Fever (>37.5°C)<br>Cough<br>Shortness of breath<br>Sore throat<br>Rhinorrhea<br>Myalgias | 24 (72.7)<br>22 (66.7)<br>21 (63.6)<br>3 (9.1)<br>3 (9.1)<br>11 (33.3) | 80 (27.6)<br>114 (39.3)<br>104 (35.9)<br>16 (5.5)<br>26 (9.0)<br>29 (10) | <0.001<br>0.003<br>0.002<br>0.41<br>0.98<br><0.001 |
| Headache | 7 (21.2) | 30 (10.3) | 0.06 |
| Change in mental status | 6 (18.2) | 42 (14.5) | 0.57 |
| **Laboratory findings** |  |  |  |
| Leukocytosis ($>11$) $\times 10^9/L$ | 2 (6.3) | 137 (47.2) | $<0.001$ |
| Lymphopenia ($\leq 1.1$) $\times 10^9/L$ | 22 (66.7) | 143 (49.7) | 0.064 |
| CRP($>3.1$) mg/L | 22 (91.7) | 177 (82.7) | 0.26 |
| **Radiological findings** |  |  |  |
| Abnormalities on CXR – no./total | 25 (83.3) | 89 (39.6) | $<0.001$ |
| Abnormalities on CT chest – no./total | 3 (100) | 39 (45.3) | 0.06 |
| **Clinical outcomes at data cutoff – no./(%)** |  |  |  |
| ICU admission | 8 (24.2) | 28 (9.7) | 0.01 |
| Mortality at 30 days | 7 (21.2) | 18 (6.2) | $\leq 0.01$ |

With regards to clinical features at presentation, PCR positive cases were more likely to have cough 66.7% vs 39.3%, shortness of breath 63.6% vs 35.9% and fever 72.7% vs 27.6%. Leukocytosis was present in 6.3% of PCR positive and 47.2% of PCR negative patients (p<0.001). Lymphopenia (≤1.1 ×10^9^/L) was present in 66.7% of PCR positive and 49.7% of PCR negative cases (p=0.064).

Positive chest x-ray (CXR) findings were reported in 83.3% of the PCR positive cases versus 39.6% of PCR negative patients (p < 0.001). Positive Chest CT features were present in 100% of PCR positive cases compared to 45.3% of PCR negative cases (p=0.062). Of the PCR positive cases, 4 (12.9%) cases were mild, 14 (45.2%) were moderate, 5 (15.2%) were severe, and 8 (25.8%) were critical; 2 (6.1%) cases were unable to be classified.

There were 24.2% and 9.7% ICU admissions in the PCR positive and negative groups, respectively (p=0.012). Mortality occurred within 30 days in 21.2% of PCR positive and 6.2% of PCR negative cases (p=0.002) (Table 1).

All patients had an NP swab collected initially. Subsequent samples included repeat NP swabs (102), sputum (6), saliva (3), tracheal aspirate (2), bronchoalveolar lavage (3) and rectal swab (1). There were no cases that initially tested negative by NP that subsequently tested positive by an alternative sampling method. In two cases, an initial negative PCR was followed by a positive PCR test occurring during the same clinical episode (within the 30 days). These cases were coded as false negatives by initial PCR.

### Clinical assessment

On clinical assessment of PCR negative cases, 245, 37, and 8 were deemed low, moderate and high probability for COVID-19, respectively. Of PCR positive cases, 1, 3 and 29 were assigned low, moderate, and high probability, respectively. Inter-assessor reliability of cases with moderate or high clinical suspicion was 68.4% (k=0.68) [18]. In cases where composite standard was available, clinical assessment of high likelihood correctly identified 91.2% of true cases, whereas of cases with moderate and low clinical likelihood, 11.5% and 0.4% were true cases, respectively.

### PCR comparison with Serology

Serology was available for 90 (27.9%) of included patients, of which 17 tested “reactive” and 73 tested “non-reactive”. Serology was done at a mean of 69 days (range 14 to 138 days) from symptom onset. The agreement between PCR and Serology was 98.9%, with one PCR positive case testing non-reactive on serology (COI=0.73) (Table 3).

### PCR comparison with composite reference standard

There were 90 patients included in the composite reference standard: 19 (21.1%) positive, 71 (78.9%) negative. Compared to the composite reference standard initial PCR was found to have no false positives and 2 false negative. Sensitivity was estimated as 94.7% (95% CI 74.0 to 99.9%), specificity was 100% (95% CI 94.9 to 100%), positive predictive value was 100% (95% CI 81.5 to 100%) and a negative predictive value was 98.6% (95% CI 92.5 to 100%) (Table 2).

**Table 2:** Diagnostic accuracy of PCR compared to composite reference standard.

| Feature | Result | Confidence interval |
| --- | --- | --- |
| Clinical Sensitivity | 94.7% | (95% CI 74.0% – 99.9%) |
| Clinical Specificity | 100% | (95% CI 94.9%-100%) |
| NPV | 98.6% | (95% CI 92.5% - 100%) |
| PPV | 100% | (95% CI 81.5%-100%) |
| Prevalence | 11.4% | (95% CI 8.1% - 15.6%) |

**Table 3:** Comparison of PCR to Serology for SARS-CoV-2 diagnosis.

|  | Serology |  |  |
| --- | --- | --- | --- |
| PCR | Positive | Negative | Total |
| Positive | 18 | 1* | 19 |
| Negative | 0 | 71 | 71 |

\*COI=0.73
Overall Percent Agreement= 98.9%, (95% CI 94.0%-99.9%)

## Discussion

In our study population, older age and male gender were associated with PCR positivity in keeping with previous reports of elderly and male patients having more severe disease requiring hospitalization [19]. Rates of epidemiological links to a known COVID-19 exposure were significantly higher in PCR positive cases at 31.4%. As previously reported, cough, fever, shortness of breath and myalgia were more frequently seen in PCR positive cases admitted to hospital [6]. Lymphopenia was present in 66.7% of PCR positive and 49.7% of PCR negative cases although this difference was not statistically significant (p=0.064). Observed trend of higher proportion of lymphopenia in PCR positive cases did not reach statistical significance perhaps as a result of high overall prevalence of lymphopenia in our study population due to other causes (HIV, solid organ transplantation, steroid therapy etc.) and small number of cases. Leukocytosis however, was significantly more frequent among PCR negative cases suggesting alternative diagnoses (i.e. bacterial causes of infection). Rates of severe outcomes including ICU admission of 24% and mortality of 21% were higher in the PCR positive group and similar to previous reports in hospitalized patients including early reports from Wuhan and more recent data from the UK [6] [20].

Positive CXR findings were more likely to be present in PCR positive cases compared to PCR negative cases (p<0.001); presence of positive CT chest finding did not reach statistical significance (p=0.095) between the two groups, perhaps due to limited numbers. Studies comparing the diagnostic accuracy of CT chest to PCR reported higher sensitivity but poor specificity (∼25%) of CT chest [21] [22] [10] [11] [23]. CT chest for primary screening or diagnosis of COVID-19 is not helpful in low prevalence setting due to significant rate of false positives [24]. Even though radiological findings may be helpful as an added diagnostic tool in settings of high COVID-19 incidence, they not only lack specificity but can also be falsely negative early after symptom onset [11] [15].

The estimated clinical sensitivity of NP swab PCR of 94.3% from this study echoes a study by Miller et al. who similarly ascertained PCR sensitivity of 95% in the first 5 days post-symptom onset in hospitalized patients [25]. Earlier studies have suggested clinical sensitivity of molecular assays to be in the range of 70% [26] [27] [28]. One of the earlier studies was a letter by Wang et al which included only 8 “nasal swabs” in their analysis [26]. While there is much interchangeable use of “nasal” and “nasopharyngeal (NP) swabs” in the literature, these are considerably different collection methods with “nasal swabs” being much less sensitive [29] [30] [31] [32]. Our group has further demonstrated the importance of proper specimen collection method with increased rate of false negative results in inadequately collected NP swabs [32]. Yang et al. (pending peer review) also reported low sensitivity of PCR, however they similarly included nasal swabs and utilized molecular assay with lower analytical sensitivity [27] [33] [34]. Although their study had limited data on patient characteristics and confirmation of COVID-19 diagnosis, it did suggest improved accuracy of lower respiratory tract specimens in diagnosis of more severe cases of COVID-19 [27]. Indeed pathogenesis of SARS-COV-2 demonstrates that the virus, once acquired, replicates in the upper respiratory tract and in a subset of patients advances and propagates in the lower respiratory airways and alveoli causing a more severe presentation [35]. However, there are practical challenges in obtaining lower respiratory tract specimens due to difficulty with sputum production or concerns of aerosol generation with bronchoscopy. In our study, only in a small proportion (3.4%) of patients had lower respiratory swabs collected.

There were 2 false negative cases on initial PCR converting to positive on subsequent testing. In the first case, the initial negative NP swab was taken 24 hrs after symptom onset and became positive 8 days later. PCR testing earlier than 48 hours of symptom onset can lead to false negative results as viral shedding can be below the level of detection [36]. The second false negative occurred in an elderly patient with severe viral pneumonia and high clinical likelihood of COVID-19. The patient’s first three NP swabs were negative until the fourth one tested positive 18 days after symptom onset. In severe infection viral load tends to be higher and peaks later; in this patient, it is possible that a lower respiratory specimen would have yielded better viral RNA recovery [37]. Clinical diagnostic accuracy of a SARS-CoV-2 PCR assay depends on the timing of presentation, clinical syndrome, anatomical site of testing, and quality of specimen collection, all of which are separate from the analytic performance of the assay itself [38] [32] [36].

As no accepted true gold standard for diagnosis of COVID-19 is available, we have developed a pragmatic composite reference model based on clinical assessment by medical experts, PCR and serology. A number of clinical prediction models have been proposed, however they have suffered from a high risk of bias and a lack of validation, therefore we opted against using them in our study [39]. Our model, although practical, is affected by the subjective nature of clinical likelihood assessment. Expert group inter-assessor reliability was 68.4% in assigning patients to moderate or high clinical likelihood groups. The moderate agreement could be reflective of non-specific nature of COVID-19 presentation as well as the risk attitudes of assessors [40]. In order to mitigate some of this subjectivity, we assigned patients deemed moderate or high likelihood category as positive on clinical assessment and cases in low likelihood category as negative. PCR and serology are incorporated as objective parts of the model; however, the evaluators were blinded to serology but not PCR results, which may have introduced bias in the clinical assessment.

Serological immunoassays have more recently become an adjunct to testing for COVID-19 [12] [41]. Similar to nucleic acid testing, serological diagnostic accuracy is determined by the target antigen and timing of collection. For example, antibodies (Abs) targeting the SARS-CoV-2 N protein are detectable earlier than antibodies against S protein, and Anti-N-protein IgG Abs tend to decrease earlier in the disease course than Anti-S-protein IgG Abs [42]. Our study criteria of serological inclusion is reflective of the fact that antibodies are more reliably detectable after two weeks and begin to decrease at four months after symptom onset [42] [12] [43] [41]. The correlation of PCR with serology was 98.9%, further increasing our confidence in the diagnostic performance on properly collected NP swabs. There were 2 false negative serology results compares to composite reference standard. While 99% of truly negative serological cases had COI value <0.1 (data not shown), one of the false negative cases had COI value of 0.73 closer to a cut-off for positivity. This case was a heart transplant recipient who was deemed a true case based on high clinical likelihood and a positive PCR. Perhaps due to his immunosuppression, this patient was unable to mount a strong immune response. Interestingly, another true COVID-19 case with prior renal transplant had a serology level at the threshold of positivity (COI=0.99) as well. The second false negative case was an elderly patient on rituximab for rheumatoid arthritis (COI=0.099). Recent studies have observed blunted or absent serological response in transplanted patients and persons taking immunosuppressive medication [44] [45]. Further work is needed in characterizing patients with low levels of IgG/IgM due various immunodeficiencies and interpretation of serological results in immunocompromised individuals should be done with caution.

The limitations of our study are the retrospective design and selection bias due to a hospital setting. Our data on clinical sensitivity applies to sicker, hospitalized patients who tend to have greater viral shedding, possibly leading to improved rates of PCR detection [46] [37]. Clinical assessment of COVID-19 likelihood was not blinded to PCR results as reviewers had access via an electronic medical record. Additionally, we have included the initial PCR test under evaluation as part of the composite reference standard. Even though we have expanded the reference standard to include any subsequent PCR results, this could have artificially enhanced the diagnostic performance of the index PCR and introduced a bias. The proposed reference standard has not been yet fully validated but reflects a practical approach utilizing currently available diagnostic modalities. The presence of a clear alternative diagnosis was used to assign lower likelihood of COVID-19 infection and although unlikely, there may have been cases of dual diagnoses. Furthermore, we were only able to obtain serology on a subset of patients as many were discharged from hospital prior to the time required to develop antibodies and had no subsequent bloodwork. Similar to PCR, false negative serology on patients could have overestimated PCR sensitivity, but in our data set, this is a rare occurrence which primarily applies to highly immunocompromised individuals.

## Conclusion

In summary, the risk of false-negative results with nucleic acid amplification tests is mostly related to pre-analytical factors such as timing of collection, the quality of sampling method and specimen type [36] [42]. Molecular testing on NP swabs has a high clinical sensitivity and excellent correlation with serology. As recommended by IDSA guidelines, cases with high clinical likelihood of COVID-19 and repeatedly negative NP swab PCR should undergo testing with serology to further enhance diagnostic yield [47], and a single PCR result cannot be interpreted in isolation without full clinical assessment of the case.

## Data Availability

Data was summarized in the Tables. Additional data needed can be made available upon request.

## Acknowledgements

The authors would like to thank Dr. Christopher Ryerson, Dr. Jane McKay and the Post-COVID-19 Recovery Clinic for their support of this work. We would like to extend our appreciation to Dr. Althea Hayden and Dr. Rohit Vijh for their guidance. We are also grateful to our medical microbiology and biochemistry technologists and scientists for their commitment to providing high quality of laboratory testing.

